# Wide-awake local anaesthesia no tourniquet hand surgery in Australia: an analysis of adoption, perceptions, and implementation in a global context

**DOI:** 10.1101/2025.11.06.25339720

**Authors:** Freya Roberts, Brett McClelland, Ali Gholamrezaei, Dion Sandoz, Elisabet Hagert, Tanya Burgess

**Affiliations:** University of Newcastle, Newcastle, NSW, Australia; Newcastle Hand Institute, Newcastle, NSW, Australia; Hunter Medical Research Institute, Newcastle, NSW, Australia; Aspetar Orthopaedic and Sports Medicine Hospital, Doha, Qatar; Qatar University, Dept of Health and Clinical Sciences, College of Medicine, Doha, Qatar; Karolinska Institute, Dept of Clinical Science and Education, Stockholm, Sweden

**Keywords:** Hand, Orthopedics, Surveys and Questionnaires, Tourniquets, Local Anaesthesia

## Abstract

**Background:** Wide-awake local anaesthesia no tourniquet (WALANT) is increasingly used globally for hand surgery, offering clinical and operational benefits. However, its adoption in Australia remains understudied. This study assessed current WALANT usage among Australian hand surgeons to identify key factors influencing its implementation.

**Methods:** A national cross-sectional survey of all Australian Hand Surgery Society (AHSS) active members (*n* 164) was conducted in 2024. The survey examined current WALANT use, perceived benefits and drawbacks, as well as facilitators and barriers of incorporation into practice. Quantitative data were analysed using descriptive and inferential statistics and qualitative data were analysed thematically.

**Results:** Sixty-eight surgeons responded (41% response rate). Seventy percent had performed WALANT at least once, and 50% incorporated it in practice. Current WALANT use was associated with hand surgery fellowship (OR 8.7), prior WALANT exposure during training (OR 6.1), and practice in university/teaching hospital (OR 3.2). Respondents highlighted avoidance of general anaesthesia, shorter recovery, improved patient satisfaction, and lower costs as benefits. Conversely, workflow inefficiencies, limited outpatient infrastructure, concerns over anaesthetist scheduling and remuneration, and the absence of dedicated billing codes were the dominant barriers. Non-users viewed WALANT as more complex and remained wary of adrenaline safety, whereas users perceived more benefits and reported few technical concerns.

**Conclusion:** WALANT is used by a substantial proportion of Australian hand surgeons, but broader adoption is limited by systemic and institutional barriers contributing to perception gap. Training, infrastructure, policy reform, appropriate funding mechanisms, and interprofessional collaboration may support wider integration of WALANT in Australia.

## INTRODUCTION

Wide-Awake Local Anaesthesia No Tourniquet (WALANT) is a technique for hand and upper extremity surgeries which utilises local haemostatic (e.g. adrenaline) and anaesthetic (e.g. lidocaine) agents to block pain and minimise bleeding, thus eliminating the need for tourniquets.^1^ Since its formal conception by Canadian hand surgeon Dr Lalonde in 2005,^2^ WALANT has become an increasingly popular technique in hand surgery, allowing for a fully ambulatory setting and encouraging patient participation during the operation, which can lead to improved surgical outcomes and better pain experience.^3^ The dynamic patient engagement allows for intraoperative assessment of motion control and immediate feedback.^4^ WALANT also offers a useful option for patients who are unsuitable for general or regional anaesthesia.^5,6^ Emerging evidence indicates the benefits of utilising WALANT for a range of hand and upper extremity surgeries and there is growing interest globally to implement this technique in routine practice.^7^ Surveys of the American Society for Surgery of the Hand (ASSH) members have shown that WALANT use is increasing among hand surgeons, though barriers still exist for widespread adoption.^8^ Lawand et al. recently conducted a systematic review on WALANT complications and side effects including 79 studies from Europe, North America, Asia, Middle East, South America, and Africa, but no published report was found originating from Australia.^9^ This raises the question of current state of WALANT practice within Australia and the need to investigate potential barriers of implementation. This cross-sectional study aims to address the knowledge gap in the current practice of WALANT in Australia and seeks to identify the factors that may influence its adoption.

## Methods

This article is prepared according to the Strengthening of The Reporting of Observational Studies in Epidemiology (STROBE) statement^10^ (STROBE checklist can be accessed in the supplementary data).

### Study design, setting, and participants

This cross-sectional survey was conducted using an online questionnaire distributed to registered members of the Australian Hand Surgery Society (AHSS), the peak professional body representing hand surgeons in Australia. Membership of the AHSS requires Fellowship of the Royal Australasian College of Surgeons (FRACS) or an equivalent qualification, along with formal training in hand surgery. Most members have backgrounds in orthopaedic or plastic surgery.

Eligible participants were surgeons currently practicing in Australia and performing hand and wrist surgeries. Survey invitations were distributed via AHSS email in September 2024, accompanied by a brief study overview and a link to the questionnaire. A reminder email was sent in December 2024, and the survey remained open for an additional three months, with a total survey time of 6 months.

### Data collection form

The survey was developed using REDCap (Research Electronic Data Capture)^11^ with items informed by a literature review on WALANT (particularly surveys in other countries),^8^ and structured according to The Consolidated Framework for Implementation Research (CFIR).^12^ The survey aimed to explore the integration of WALANT into hand surgery practice in Australia, with the goal of identifying barriers and opportunities for improvement and informing future implementation strategies. The questionnaire was anonymous, concise, and designed to be user-friendly, requiring no more than five minutes to complete. It was divided into three sections: (1) current practice of WALANT, (2) perspectives on WALANT, including perceived barriers and facilitators, and (3) Surgeon and practice profile (Table 1). A copy of the survey instrument is provided in the supplementary data.

**Table 1.** Overview of survey content.

| **Domain** | **Key areas assessed** |
| --- | --- |
| **Current WALANT practice** | Prior and current use; years of experience; types of procedures and frequency of WALANT use for each procedure; settings; who administers anaesthetic; reasons for non-use |
| **Perspectives on WALANT** | Perceived complexity and ease of implementation; influence of system, organisational, and individual factors; perceived benefits; perceived drawbacks; free-text on facilitators and barriers |
| **Surgeon and practice profile** | Role and specialty; fellowship training; work settings; years of practice; annual surgical volume; proportion of hand/wrist cases; practice population and location |

### Study size and Bias

The invitation was sent to all registered members of the AHSS (i.e., a census), thereby minimising coverage bias. Based on a response rate of approximately 50% reported in similar surveys ^13, 14^ we anticipated receiving around 82 usable responses from 164 active members of the AHSS. A sample size of 82 would have provided a margin of error of ±7 percentage points when estimating a proportion of 50% (i.e., a conservative estimate). However, we received 68 eligible responses, yielding an achieved margin of error of ±9 percentage points around an observed proportion of 51% (see Results). While this is less precise than anticipated, it remains adequate for the descriptive objectives of the study.

### Analysis

Quantitative data were analysed using JASP (version 0.17.3, University of Amsterdam, Netherlands) employing both descriptive and inferential statistics. Results are presented as number (percentage) for categorical variables and median [interquartile range] for ordinal and continuous variables. The Chi-Square Test (or Fisher’s Exact Test) and Mann-Whitney U Test were used to examine associations with the current practice of WALANT. A *P* value of <0.05 was considered statistically significant.

Qualitative data from free-text responses were analysed using thematic analysis,^15^ assisting identification of key patterns and further insights related to barriers, facilitators, and perceptions of WALANT implementation.

### Ethics Approval

This study received ethics approval from the College Human Ethics Advisory Panel, University of Newcastle (approval number H-2024-0225), in accordance with the provisions of the National Statement on Ethical Conduct in Human Research.^16^

## Results

### Participant characteristics

A total of 68 active AHSS members completed the survey, yielding a response rate of 41%. Respondents represented a diverse range of practice locations across all Australian states except the Northern Territory, with the majority practising in New South Wales (27, 40%), Victoria (16, 23%), and Western Australia (9, 13%).

Most participants were practising in private settings (62, 91%) and/or university hospitals (39, 57%), with metropolitan populations comprising the main catchment area for 51 (75%) respondents. Approximately half had been in practice for over 20 years (32, 47%) and 42 (62%) reported performing more than 500 surgeries per year on average.

In terms of specialty, 44 participants (65%) were trained in orthopaedic surgery, and 24 (35%) in plastic surgery. The majority (60, 88%) had completed a fellowship in hand surgery. For 49 participants (72%), hand and wrist surgery accounted for more than 50% of their clinical practice (refer to Table S1 in Supplementary Data for additional details).

### WALANT practice

Out of 68 participants, 48 (71%) reported having ever used WALANT, and 35 (51%) were currently using WALANT in their clinical practice. Among current users, 66% (23/35) had been using WALANT for more than five years, and the majority (86%, 30/35) reported using it primarily in inpatient hospital settings. Additionally, 31% (11/35) utilised WALANT in outpatient clinics, with 14% (5/35) performing WALANT exclusively in the outpatient setting. Most current users (91%, 32/35) reported personally administering the local anaesthetic during WALANT procedures.

### Common procedures for using WALANT

The most performed procedures using WALANT were elective tendon repairs (97%, 34/35) and elective nerve procedures (80%, 28/35). WALANT was less frequently used for arthroplasty (11%, 4/35) and not used at all for wrist fractures. The range of procedures and the proportion of patients treated with WALANT for each procedure are shown in Figure 1.

### Potential barriers and facilitators to using WALANT

Participants not currently using WALANT (*n* = 33, including those who had never used it) were asked to indicate their reason(s), using pre-defined options and free-text responses. The most frequently cited barriers were related to workflow inefficiency and operational challenges (e.g., visualisation issues), followed by knowledge and acceptance gaps (e.g., lack of training, low patient preference), and clinical safety concerns (e.g., use of adrenaline).

Thematic analysis of free-text responses identified additional themes, including concerns about disrupting established surgeon-anaesthetist workflow and uncertainty regarding anaesthetist or team remuneration under the WALANT model (see Figure 2 and Table S6 and S7 in Supplementary File for detailed findings).

All participants, regardless of whether they currently use WALANT, were asked to rate the factors influencing their decision to use or not use it. While most respondents rated the pre-defined factors as having no or little influence, several individual (e.g., knowledge, skills, attitudes), practice-level (e.g., culture, competing priorities), and organisational (e.g., support, resources) factors were rated as having moderate to strong influence by approximately 30-40% of the participants (Figure 3).

Participants were also asked about their perspectives on the potential benefits and drawbacks of WALANT. While most respondents (> 65%) agreed that WALANT enables patient feedback, reduces general anaesthesia risks, and is convenient for outpatient procedures, there was greater uncertainty or disagreement regarding its impact on surgical time, recovery, and clinical outcomes (Figure 4). The most endorsed drawbacks were injection pain, psychological stress, and patient anxiety/discomfort. In contrast, concerns about toxicity, anaesthetic failure, and limited applicability were less widely agreed upon (Figure 5).

### Processes supporting WALANT implementation

Qualitative analysis of responses regarding processes supporting WALANT implementation and factors facilitating its adoption in Australia revealed several key themes (Table S6 and S7 in Supplementary Data). Participants described WALANT uptake as being influenced primarily by system-level factors, rather than technical considerations. The most frequently cited barriers included (1) anaesthetist availability and remuneration, (2) Medicare/insurance and hospital billing rules, and (3) lack of procedure-room space, all of which disrupt theatre list organisation. Cultural expectations, among both patients and staff, for general anaesthesia were also noted, though perceived as secondary. Reported facilitators included access to protected WALANT lists or outpatient rooms, early patient education, and explicit support from theatre teams and management.

### Factors associated with WALANT use

Participants who currently used WALANT were compared to those who did not in terms of practice profile and perspectives toward WALANT. WALANT users were significantly more likely to have completed a fellowship in hand surgery (OR = 9.1, 95% CI: 1.0–79.1, p = 0.044), received specific training in WALANT (OR = 6.2, 95% CI: 1.2–30.9, p = 0.026), practice in a teaching hospital (OR = 5.1, 95% CI: 1.9–13.6, p < 0.001), and serve primarily metropolitan populations (OR = 3.4, 95% CI: 1.0–11.1, p = 0.041).

No significant differences were observed between participants who currently use WALANT and those who do not across any of the factors influencing the decision to use or not use it (Figure S3 in Supplementary Data). Those who currently use WALANT were more agreeable to its potential benefits such as less general anaesthesia risk (p = 0.034), more patient comfort (p < 0.001), awake patient feedback (p = 0.012), cost savings (p < 0.001), faster recovery (p < 0.001), shorter surgery time (p < 0.001), better outcomes (p < 0.001), outpatient convenience (p = 0.003), and fewer complications (p = 0.008) (Figure S4 in Supplementary Data). They were also less likely to agree with common concerns such as incomplete anaesthesia (p = 0.005), time-consuming injection (p = 0.006), difficult haemostasis (p = 0.025), and local anaesthetic toxicity (p <0.001) (Figure S5 in Supplementary Data).

## Discussion

This survey provides the first national data on WALANT adoption in Australia, showing that 70% of respondents have used the technique and 51% incorporate it regularly. The typical WALANT user was fellowship-trained (often in orthopaedics) and practicing in a metropolitan teaching hospital, suggesting that early adopters tend to be highly trained, academically connected, and working in high-volume, resource-rich settings. Indeed, our analysis found that prior WALANT training and teaching-hospital practice were strong predictors of use (OR 6.2 and OR 5.1, respectively), implying that uptake is closely tied to structured education and institutional support rather than being purely grassroots. These findings align with international observations. In an American Society for Surgery of the Hand (ASSH) survey, 79% of members had tried WALANT and 62% currently used it.^8^ Surgeons exposed to WALANT during training were far more likely to adopt it (87% vs 63%),^8^ and those in early-career or very high-volume practices also reported higher use (for example, 75% of surgeons with ≤10 years in practice vs 58% of those >20 years used WALANT.^8^ Likewise, recent reviews note that WALANT is safe and increasingly popular across a wide range of hand procedures.^17^ Our results mirror global trends showing that WALANT integration depends on training and practice context.

### The perception gap and systemic barriers

Despite its technical feasibility, we found a disconnect between WALANT’s potential benefits and real-world practice. Many respondents perform WALANT for routine tendon and nerve releases, but the main reasons for not using WALANT more widely were systemic rather than clinical. Workflow inefficiency was cited by 48% of non-users, and qualitative responses emphasised obstacles like anaesthetist scheduling, absence of specific outpatient billing codes, and rigid theatre allocation. In effect, surgeons who have developed workarounds (e.g. dedicated WALANT clinic lists) tend to find the technique highly advantageous, whereas others perceive it as onerous to implement. This pattern echoes international reports: for example, in an ASSH survey 51% of surgeons still required anaesthesia support for WALANT cases and only about 2% had concerns about epinephrine use.^8^ Likewise, lack of familiarity, operating-room efficiency concerns, and patient preferences were identified as substantial barriers to WALANT adoption in ASSH survey.^8^ In our survey too, administrative and logistical factors, such as procedure location and workflow flexibility, emerged as the primary hurdles to broader use.

### Clinical and policy implications

These barriers have clear implications for practice and health policy. First, the strong link between WALANT training and use indicates a need to build WALANT into surgical education. Residency and fellowship programs in hand surgery can include hands-on WALANT experience, along with training in patient communication and efficient clinic workflows. Indeed, international data show that surgeons who learn WALANT in training are significantly more likely to use it in practice.^8^

Second, financial and regulatory reforms could incentivise shifting appropriate cases out of main theatres. For example, establishing dedicated Medicare item numbers or funding pathways for clinic-based WALANT procedures could help overcome the current disincentive to operate outside the operating room. Such a shift is supported by evidence of substantial cost and time savings: a study of trigger-finger release found that WALANT in a procedure room was markedly cheaper and faster than the identical surgery in a traditional OR.^2^

Third, at the institutional level hospitals can facilitate adoption by creating WALANT-friendly infrastructure. Several respondents noted success with dedicated procedure rooms and lists for WALANT cases; formalising these models would uncouple straightforward hand cases from anaesthetist-dependent theatres. Importantly, broader use of WALANT has been associated elsewhere with improved service delivery, for instance, trauma units reported better time-to-treatment performance, higher patient satisfaction, and lower costs when using WALANT.^18^ These findings suggest that policy changes and support for WALANT programs could enhance efficiency without compromising outcomes.

### Strengths and limitations

Our study benefits from its national scope, surveying the entire membership of the AHSS, and from a mixed-methods design that captured both usage statistics and qualitative insights. Anonymity likely encouraged frank feedback on sensitive topics like interprofessional relations and reimbursement. However, the 41% response rate introduces potential bias: surgeons with strong opinions about WALANT (either advocates or sceptics) may have been more likely to respond. Consequently, our finding that 51% of respondents are current users may overestimate the true prevalence among all Australian hand surgeons. For context, a recent ASSH survey had a 23% response rate.^8^ Thus, these results should be interpreted as reflecting the engaged early adopters and their perspectives, rather than a definitive census. Finally, this cross-sectional design provides a snapshot in time and cannot capture trends in adoption.

### Summary

WALANT use is common in Australia but remains uneven, with adoption driven more by logistical feasibility than by doubts about clinical benefits. Key enablers include improved workflow integration, supportive institutional policies, dedicated outpatient setups, and inclusion of WALANT in surgical training. Collaborating with anaesthetists in evolving care models is also vital to maintain professional cohesion.

While WALANT is well supported for safety and efficacy, further research is needed to evaluate its system-level impact, cost-effectiveness, patient experience, and implementation in diverse settings. Future studies can explore its role in trauma pathways, rural care, and elective surgery backlogs. The findings of the present study offer a foundation for national guidelines to support broader, evidence-based integration of WALANT into Australian surgical practice.

## Data Availability

All data produced in the present study are available upon reasonable request to the authors

## Acknowledgements

We would like to express our appreciation for the support of the Australian Hand Surgery Society in facilitating the survey. The Newcastle Hand Institute provided in-kind support, including staff time, for conducting the survey and preparing this report.

## Data Sharing and Data Accessibility

Data is available from the Corresponding Author upon reasonable request.

## Author Contributions

TB, BM, DS, AG, and EH conceptualised the study and developed the study protocol and materials. AG collected the data. AG and FR analysed the data. All authors contributed to interpreting the results. AG and FR drafted the manuscript, and all authors contributed to revising it critically for important intellectual content.

## Figure Legends

Figure 1: Procedures and percentage of patients for whom WALANT is used. Shading indicates usage intensity by patient volume (n = 35 current WALANT users)

Figure 2: Reason(s) for not using WALANT among respondents (*n* = 33)

Figure 3: Factor influencing the decision to use or not use WALANT (*n* = 68)

Figure 4: Perspective toward potential benefits of using WALANT (*n* = 68)

Figure 5: Perspective toward potential drawbacks of using WALANT (*n* = 68)

## List of Supporting Information

Appendix A: Supplementary Data

Appendix B: Survey Questionnaire

Appendix C: STROBE statement

