## Supplementary Data for "Wide-awake local anaesthesia no tourniquet hand surgery in Australia: an analysis of adoption, perceptions, and implementation in a global context"

**Appendix A: Supplementary Data**

**Table S1: Characteristics of the respondents (*n* 68)**

| | | Frequency | Valid Percent |
|---|---|---|---|
| ***Current job title*** | | | |
| *(Multiple choice)* | Staff Specialist | 10 | 14.7 |
| | Visiting Medical Officer | 43 | 63.2 |
| | Private Practice | 51 | 75 |
| | Missing | 0 | |
| ***Surgical specialty*** | | | |
| | Orthopaedic Surgeon | 44 | 64.7 |
| | Plastic Surgeon | 24 | 35.2 |
| | Missing | 0 | |
| ***Current place(s) of work?*** | | | |
| *(Multiple choice)* | Teaching / University Academic Hospital | 39 | 57.3 |
| | Non-Academic Public Hospital | 12 | 17.6 |
| | Private Practice | 62 | 91.1 |
| | Missing | 0 | |
| ***Fellowship in Hand Surgery*** | | | |
| | YES/NO | 60/8 | 88.2/11.7 |
| | Missing | 0 | |
| ***Training in WALANT*** | | | |
| | YES/NO | 12/55 | 17.9/82.0 |
| | Missing | 1 | |
| ***Duration of practice*** | | | |
| | < 5 years | 10 | 17.7 |
| | 5-10 years | 4 | 5.8 |
| | 11-20 years | 22 | 32.3 |
| | >20 years | 32 | 47.0 |
| | Missing | 0 | |
| ***Number of surgeries performing each year*** | | | |
| | < 200 | 3 | 4.4 |
| | 200-500 | 23 | 33.8 |
| | > 500 | 42 | 61.7 |
| | Missing | 0 | |
| ***% of practice involving hand and wrist surgery*** | | | |
| | 0-25 % | 1 | 1.4 |
| | 26-50 % | 18 | 26.4 |
| | 51-75 % | 17 | 25.0 |
| | 76-100 % | 32 | 47.0 |
| | Missing | 0 | |
| ***Main population in practice*** | | | |
| | Metropolitan | 51 | 75.0 |
| | Rural/Regional | 17 | 25.0 |
| | Missing | 0 | |

**Table S2: WALANT practice:**

| | | Frequency | Valid Percent |
|---|---|---|---|
| ***Ever used WALANT*** | | | |
| *(total of 68)* | YES/NO | 48*/20 | 70.5/29.4 |
| | Missing | 0 | |
| ***Currently using WALANT*** | | | |
| *(*total of 48)* | YES/NO | 35**/12 | 74.4/25.5 |
| | Missing | 1 | |
| ***Years have been using WALANT*** | | | |
| *(**total of 35)* | < 2 years | 2 | 5.7 |
| | 2-5 years | 10 | 28.5 |
| | > 5 years | 23 | 65.7 |
| | Missing | 0 | |
| ***Procedures using WALANT for*** | | | |
| *(Multiple choice)* | Elective tendon procedures (i.e. Trigger finger, de Quervain) | 34 | 97.1 |
| | Acute tendon procedures (i.e. Flexor/extensor tendon repairs) | 12 | 34.3 |
| | Elective nerve procedures (i.e. Carpal/cubital tunnel release) | 28 | 80.0 |
| | Acute nerve procedures (i.e. Digital nerve repair) | 11 | 31.4 |
| | Elective soft tissue procedures (i.e. Dupuytrens, ganglion cyst removal) | 14 | 40.0 |
| | Arthroplasty procedures (i.e. CMC/MCP/IP joint) | 4 | 11.4 |
| | Fracture surgery / digits (i.e. Phalanx fracture) | 10 | 28.6 |
| | Fracture surgery / wrist (i.e. Distal radius) | 0 | 0 |
| | Missing | 0 | |
| *(Other, textual answer)* | Tendon procedures (Tenolysis, Tendon transfers, Contracture release) | 3 | - |
| | Percutaneous needle fasciotomy | 1 | - |
| ***Settings using WALANT*** | | | |
| *(Multiple choice)* | Outpatient clinic / surgery centre | 11 | 31.4 |
| | Inpatient hospital | 30 | 85.7 |
| | Missing | 0 | |
| ***Person giving local anaesthetic for WALANT*** | | | |
| *(Multiple choice)* | Surgeon | 32 | 91.4 |
| | Assistant | 2 | 5.7 |
| | Anaesthetist | 12 | 34.3 |
| | Missing | 0 | |

**Table S3: Those who selected each procedure, for what % of the patients they have you been using WALANT?**

| | ***% of patients, n (valid %)*** | | | | | |
|---|---|---|---|---|---|---|
| ***Procedure*** | **<= 10%** | **10-50%** | **50-90%** | **>= 90%** | **Total** | **Missing** |
| Elective tendon procedures (i.e. Trigger finger, de Quervain) | 8 (25.0) | 8 (25.0) | 8 (25.0) | 8 (25.0) | 32 (91.4) | 0 |
| Acute tendon procedures (i.e. Flexor/extensor tendon repairs) | 3 (25.0) | 6 (50.0) | 2 (16.7) | 1 (8.3) | 12 (34.3) | 0 |
| Elective nerve procedures (i.e. Carpal/cubital tunnel release) | 6 (22.2) | 7 (25.9) | 6 (22.2) | 8 (29.6) | 27 (77.1) | 0 |
| Acute nerve procedures (i.e. Digital nerve repair) | 4 (36.4) | 1 (9.1) | 5 (45.5) | 1 (9.1) | 11 (31.4) | 0 |
| Elective soft tissue procedures (i.e. Dupuytren's, ganglion cyst removal) | 4 (30.8) | 7 (53.8) | 1 (7.7) | 1 (7.7) | 13 (37.1) | 0 |
| Arthroplasty procedures (i.e. CMC/MCP/IP joint) | 2 (50.0) | 1 (25.0) | 0 (0) | 1 (25.0) | 4 (11.4) | 0 |
| Fracture surgery / digits (i.e. Phalanx fracture) | 4 (40.0) | 3 (30.0) | 1 (10.0) | 2 (20.0) | 10 (28.6) | 0 |

**Table S4: Reasons for not using WALANT (20 have never used, 12 not currently using, total = 33)**

| | Frequency | Valid Percent |
|---|---|---|
| ***Answer choices*** | | |
| Visualization issues | 8 | 24.2 |
| Not familiar with WALANT / Not trained | 6 | 18.2 |
| Not efficient in my practice | 16 | 48.5 |
| Not preferred by patients in my practice | 7 | 21.2 |
| Administrative barriers | 5 | 15.2 |
| Anaesthesia concerns | 6 | 18.2 |
| Outside the scope of my practice | 0 | 0.0 |
| Financial concerns | 4 | 12.1 |
| Concerns with using adrenaline (epinephrine) | 7 | 21.2 |
| Concerns regarding WALANT effectiveness | 1 | 3.0 |
| Concerns with sterility | 0 | 0 |
| Missing | 0 | |
| ***Other reasons*** *(Thematic analysis of textual answers)* | | |
| Concern that a switch would disrupt established surgeon-anaesthetist workflow or relationships | 3 | - |
| No compelling reason to change existing practices | 2 | - |
| Perception that the technique is slower or ties up consulting-room time | 2 | - |
| Doubt about how anaesthetists (or the team) would be remunerated under the new model | 1 | - |
| Personal comfort with familiar operative tools | 1 | - |

**Table S5: Factors associated with use of WALANT**

| | **Currently WALANT use** | | **OR (95% CI)** | **P** |
|---|---|---|---|---|
| **Demographic characteristics** | NO<br>n = 33 | YES<br>n = 35 | | |
| Fellowship in Hand Surgery | 26 (78.8) | 34 (97.1) | 9.1 (1.0–79.1) | 0.044 |
| Training in WALANT | 2 (6.1) | 10 (28.76) | 6.2 (1.2–30.9) | 0.026 |
| Practice in Teaching Hospital | 14 (42.4) | 25 (71.4) | 5.1 (1.9–13.6) | < 0.001 |
| Main population in practice (Metropolitan) | 21 (63.6) | 30 (85.7) | 3.4 (1.0 – 11.1) | 0.041 |

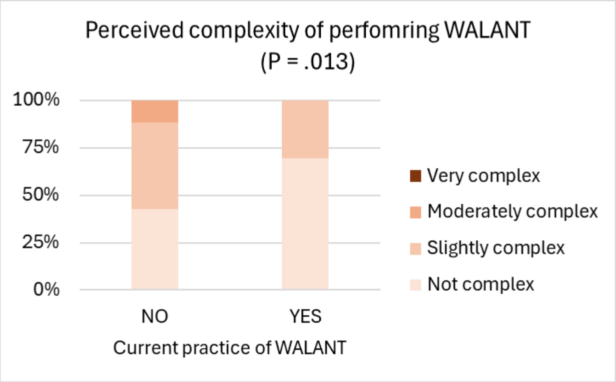


Figure S1: Perceived complexity of performing WALANT among current users and non-users (n = 68). Participants who did not currently use WALANT were significantly more likely to perceive it as complex to perform, with higher proportions rating it as “moderately” or “very” complex (p = 0.013).

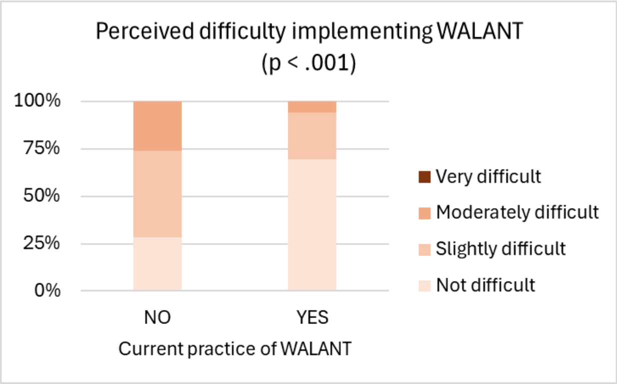


Figure S2: Perceived difficulty of implementing WALANT among current users and non-users (n = 68). Participants who did not currently use WALANT were significantly more likely to rate its implementation as difficult, with higher proportions selecting “moderately” or “very” difficult ($p < 0.001$).

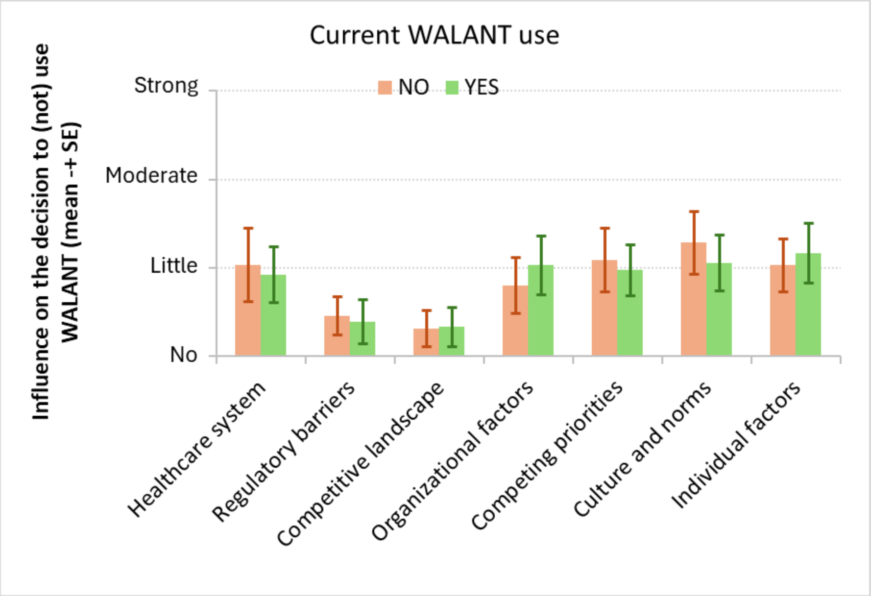


Figure S3: Perceived influence of contextual factors on the decision to use or not use WALANT, by current use status (mean ± SE, n = 68). Participants rated the influence of various factors (e.g., healthcare system, culture, individual attitudes) on a 4-point scale from "no" to "strong" influence. While some trends differed between current WALANT users and non-users, no statistically significant differences were observed across groups.

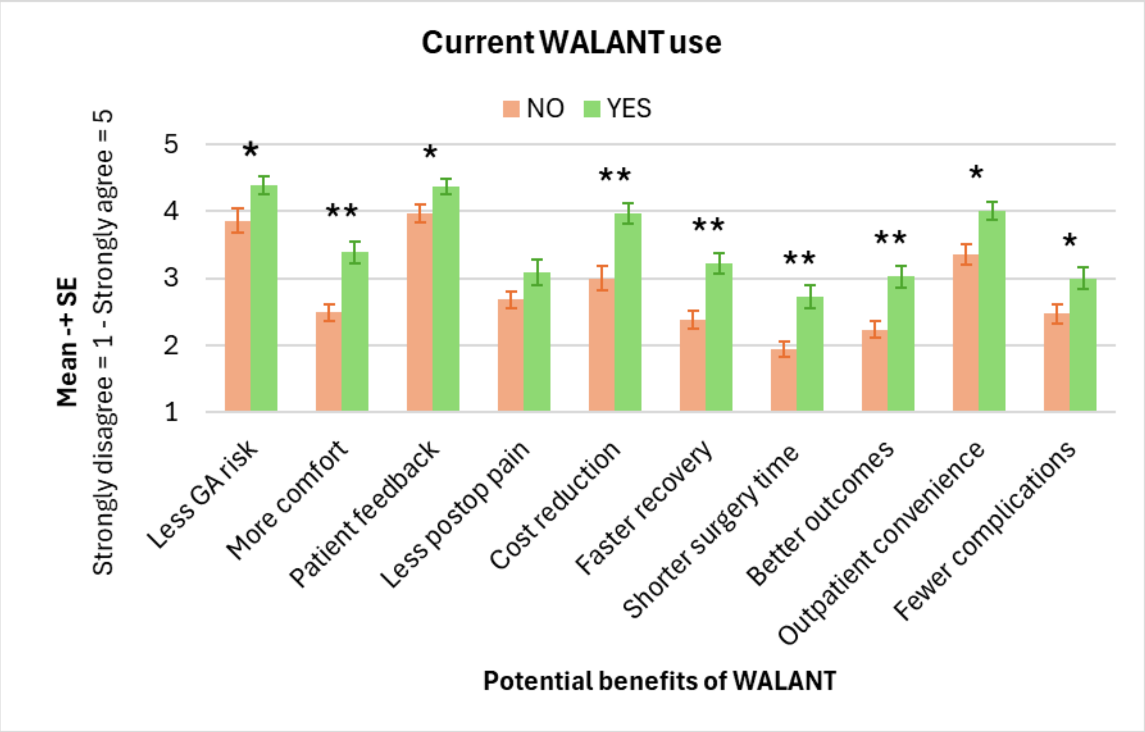

Figure S4: Those who currently use WALANT were more agreeable to its potential benefits such as less general anaesthesia risk, more patient comfort, awake patient feedback, cost savings, faster recovery, shorter surgery time, better outcomes, outpatient convenience, and fewer complications. * $p < 0.05$, ** $p < 0.001$.

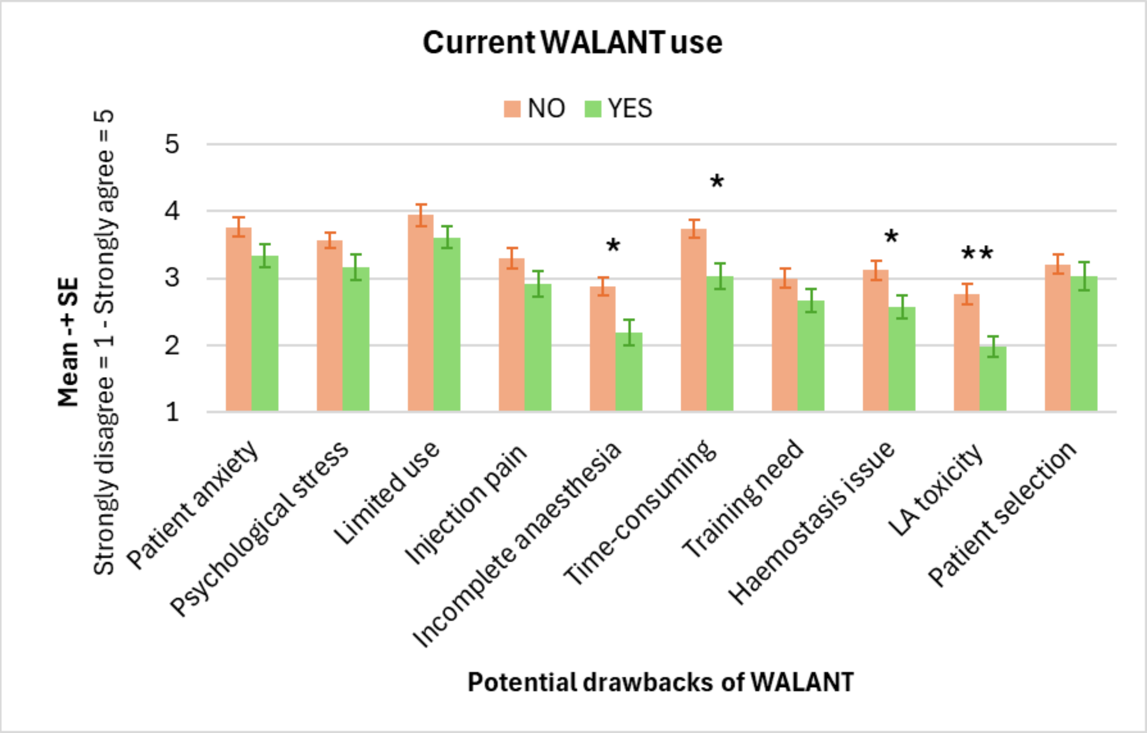


Figure S5: Those who currently use WALANT were less likely to agree with common concerns such as incomplete anaesthesia, time-consuming injection, difficult haemostasis, and local anaesthetic toxicity. * $p < 0.05$, ** $p < 0.001$.

### Appendix B: Survey Questionnaire

**[Current practice of WALANT]**

**1. Have you ever used WALANT in your practice?**

◯ YES ◯ NO

**2. [if 1 is YES] Are you currently using WALANT in your practice?**

◯ YES ◯ NO

**2.1. [If 2 is YES] How many years have you been using WALANT?**

◯ < 2 y ◯ 3-5 y ◯ > 5 y

**2.2. [If 2 is YES] For which procedures, do you use WALANT? Select all that apply**

◯ Elective tendon procedures (i.e. Trigger finger, de Quervain)
◯ Acute tendon procedures (i.e. Flexor/extensor tendon repairs)
◯ Elective nerve procedures (i.e. Carpal/cubital tunnel release)
◯ Acute nerve procedures (i.e. Digital nerve repair)
◯ Elective soft tissue procedures (i.e. Dupuytren's, ganglion cyst removal)
◯ Arthroplasty procedures (i.e. CMC/MCP/IP joint)
◯ Fracture surgery / digits (i.e. Phalanx fracture)
◯ Fracture surgery / wrist (i.e. Distal radius)
[if selected] for what % of patients? >= 90% 50-90% 10-50% <= 10%
◯ Other surgeries [if selected] Please mention:

**2.3. [If 2 is YES] In which settings have you been using WALANT? Select all that apply**

◯ Outpatient clinic / surgery centre ◯ Inpatient hospital

**2.4. [If 2 is YES] Who gives the local anaesthetic for WALANT in your practice? Select all that apply**

◯ Myself ◯ My assistant ◯ My anaesthetist

**[if 1 or 2 is NO] What are the reasons for not using WALANT in your practice? Select all that apply**

- Visualization issues
- Not familiar with WALANT / Not trained
- Not efficient in my practice
- Not preferred by patients in my practice
- Administrative barriers
- Anaesthesia concerns
- Outside the scope of my practice
- Financial concerns
- Concerns with using adrenaline (epinephrine)
- Concerns regarding WALANT effectiveness
- Concerns with sterility
- Other reasons: please specify …

**[Demographics]**

**1. What is your current job title? Please select all that apply**

- Staff Specialist
- Visiting Medical Officer
- Private Practice

**2. What is your surgical specialty?**

- Orthopaedic Surgeon
- Plastic Surgeon
- General Surgeon

**3. Have you undertaken a fellowship in Hand Surgery?**

- YES
- NO

**4. Was WALANT part of your residency or fellowship training?**

- YES
- NO

**5. What is your current place(s) of work? Select all that apply**

- Teaching / University Academic Hospital
- Non-Academic Public Hospital
- Private Practice

**6. How many years have you been practicing as a surgeon?**

◯ <5 y ◯ 5-10 y ◯ 11-20 y ◯ >20 y

**7. How many surgeries on average do you perform each year?**
◯ < 200 ◯ 200-500 ◯ > 500

**8. What percentage of your practice involves hand and wrist surgery?**
◯ 0-25% ◯ 26-50 % ◯ 51-75% ◯ 76-100%.

**9. Which is the main population in your practice?**
◯ Metropolitan ◯ Rural/Regional ◯ Remote

**10. In which state do you practice?**
◯ VIC ◯ NSW ◯ QLD ◯ TAS ◯ SA ◯ NT ◯ WA ◯ ACT

**[Barriers and Facilitators]**

**[A. Intervention Characteristics]**

**1. How would you rate the complexity of WALANT?**
◯ Not at all complex------------------------◯ Very complex [5 point Likert]

**2. How easy is it, or would it be, to implement WALANT in your practice?**
◯ Not at all easy------------------------◯ Very easy [5 point Likert]

**[C-E. Outer Setting / Inner Setting / Characteristics of Individuals]**

**1. How much does each of the following factors influence your decision to use or not to use WALANT in your practice?** [From Not at all to Very much, 5-point Likert]
a. The wider healthcare system
b. Regulatory or legal barriers
c. Competitive landscape of the practice environment
d. Organizational factors (e.g., leadership support, availability of resources)
e. Competing demands and priorities within my practice
f. Culture and norms within my practice
g Individual factors (e.g., knowledge, skills, and attitudes)

**2. Do you agree or disagree with the following potential benefits of using WALANT?**
[From Completely disagree to Completely agree, 5-point Likert]
a. Reduces risks associated with general anaesthesia
b. Increases patient comfort

| |
|---|
| c. Allows the patient to be awake and provide feedback<br>d. Reduced postoperative pain<br>e. Reduces the costs<br>f. Shorter Recovery Time<br>g. Decreased Surgical Time<br>h. Improved Clinical Outcomes<br>i. Convenience for Outpatient Procedures<br>j. Lower Risk of Complications |
| **3. Do you agree or disagree with the following potential drawbacks of using WALANT?**<br>[From Completely disagree to Completely agree, 5-point Likert]<br>a. Patient Anxiety and Discomfort<br>b. Psychological Stress<br>c. Limited Applicability<br>d. Injection Pain<br>e. Incomplete Anaesthesia<br>f. Time-Consuming Injection Process<br>g. Surgeon and Staff Training<br>h. Homeostasis Management<br>i. Local Anaesthetic Toxicity<br>j. Patient Selection |
| **[F. Process]** |
| **1. What processes are in place to support the implementation of WALANT in your practice?**<br>**Please write your answer …** |

**Are there any other factors that may be relevant to the adoption of the technique among hand surgeons in Australia?**
**Please write your answer …**

**Appendix C:** STROBE Statement—Checklist of items that should be included in reports of ***cross-sectional studies***

| | **Item No** | **Recommendation** | **Page No** |
|---|---|---|---|
| **Title and abstract** | 1 | (*a*) Indicate the study's design with a commonly used term in the title or the abstract | 1 |
| | | (*b*) Provide in the abstract an informative and balanced summary of what was done and what was found | 3 |
| | | **Introduction** | |
| Background/rationale | 2 | Explain the scientific background and rationale for the investigation being reported | 5 |
| Objectives | 3 | State specific objectives, including any prespecified hypotheses | 5 |
| | | **Methods** | |
| Study design | 4 | Present key elements of study design early in the paper | 6 |
| Setting | 5 | Describe the setting, locations, and relevant dates, including periods of recruitment, exposure, follow-up, and data collection | 6 |
| Participants | 6 | (*a*) Give the eligibility criteria, and the sources and methods of selection of participants | 6 |
| Variables | 7 | Clearly define all outcomes, exposures, predictors, potential confounders, and effect modifiers. Give diagnostic criteria, if applicable | 6-7 |
| Data sources/ measurement | 8 | For each variable of interest, give sources of data and details of methods of assessment (measurement). Describe comparability of assessment methods if there is more than one group | 7 |
| Bias | 9 | Describe any efforts to address potential sources of bias | 7 |
| Study size | 10 | Explain how the study size was arrived at | 7 |
| Quantitative variables | 11 | Explain how quantitative variables were handled in the analyses. If applicable, describe which groupings were chosen and why | 7 |
| Statistical methods | 12 | (*a*) Describe all statistical methods, including those used to control for confounding | 7 |
| | | (*b*) Describe any methods used to examine subgroups and interactions | |
| | | (*c*) Explain how missing data were addressed | |
| | | (*d*) If applicable, describe analytical methods taking account of sampling strategy | |
| | | (*e*) Describe any sensitivity analyses | |
| | | **Results** | |
| Participants | 13 | (a) Report numbers of individuals at each stage of study—eg numbers potentially eligible, examined for eligibility, confirmed eligible, included in the study, completing follow-up, and analysed | 8 |
| | | (b) Give reasons for non-participation at each stage | NA |
| | | (c) Consider use of a flow diagram | NA |
| Descriptive data | 14 | (a) Give characteristics of study participants (eg demographic, clinical, social) and information on exposures and potential confounders | 8 |
| | | (b) Indicate number of participants with missing data for each variable of interest | NA |

| | | | |
|---|---|---|---|
| Outcome data | 15 | Report numbers of outcome events or summary measures | 8-11 |
| Main results | 16 | (*a*) Give unadjusted estimates and, if applicable, confounder-adjusted estimates and their precision (eg, 95% confidence interval). Make clear which confounders were adjusted for and why they were included | NA |
| | | (*b*) Report category boundaries when continuous variables were categorized | NA |
| | | (*c*) If relevant, consider translating estimates of relative risk into absolute risk for a meaningful time period | NA |
| Other analyses | 17 | Report other analyses done—eg analyses of subgroups and interactions, and sensitivity analyses | NA |
| **Discussion** | | | |
| Key results | 18 | Summarise key results with reference to study objectives | 11 |
| Limitations | 19 | Discuss limitations of the study, taking into account sources of potential bias or imprecision. Discuss both direction and magnitude of any potential bias | 13 |
| Interpretation | 20 | Give a cautious overall interpretation of results considering objectives, limitations, multiplicity of analyses, results from similar studies, and other relevant evidence | 12-13 |
| Generalisability | 21 | Discuss the generalisability (external validity) of the study results | 14 |
| **Other information** | | | |
| Funding | 22 | Give the source of funding and the role of the funders for the present study and, if applicable, for the original study on which the present article is based | 14 |
